# Pharmacokinetics and pharmacogenomics of clozapine in an ancestrally diverse sample: A longitudinal analysis and GWAS using clinical monitoring data from the UK

**DOI:** 10.1101/2022.09.23.22280299

**Authors:** Antonio F. Pardiñas, Djenifer B. Kappel, Milly Roberts, Francesca Tipple, Lisa M. Shitomi-Jones, Adrian King, John Jansen, Marinka Helthuis, Michael J. Owen, Michael C. O’Donovan, James T.R. Walters

## Abstract

**Background:** The antipsychotic clozapine is the only drug with proven effectiveness against the treatment-resistant symptoms that affect 20-30% of those with schizophrenia. Despite this, clozapine is markedly under-prescribed, partly due to concerns about its narrow therapeutic range and adverse drug reaction profile. Both concerns are linked to drug metabolism, which varies across worldwide populations and is partially genetically determined. There is, however, a lack of clozapine pharmacogenomic data based on study participants of multiple ancestries.

**Methods:** We analysed data from 4,495 individuals linked to 16,068 assays from a clozapine monitoring service in the UK. Genomic information was used to identify five biogeographical ancestries (European, Sub-Saharan African, North African, Southwest Asian and East Asian) as well as admixed individuals. Pharmacokinetic modelling, GWAS, and a polygenic score association analysis were conducted on this longitudinal dataset using three outcome variables: two metabolite plasma concentrations (clozapine and norclozapine) and their ratio.

**Findings:** A faster average clozapine metabolism was seen in those of Sub-Saharan African ancestry compared to Europeans. In contrast, East and Southwest Asians were more likely to be slow clozapine metabolisers. Eight pharmacogenomic loci were identified in the GWAS, with consistent cross-ancestral effects. Polygenic scores generated from these loci led to significant associations with clozapine outcome variables in the whole sample and within individual ancestries, with variances explained between 0.61%-7.26%.

**Interpretation:** Longitudinal cross-ancestry GWAS can discover pharmacogenomic markers of clozapine metabolism that, individually or as polygenic scores, have consistent effects across ancestries. While the potential clinical role of these predictors is evaluated, we provide strong evidence that ancestral differences in clozapine metabolism should be incorporated into clozapine dosing and managing protocols to optimise their utility for diverse populations.

**Funding:** Medical Research Council (MRC).

## INTRODUCTION

Schizophrenia is typically a chronic disorder with symptoms that are severe and distressing for most people. The majority of individuals obtain some benefit from antipsychotic medication^1^, although between 20-30% experience treatment-resistant schizophrenia (TRS), defined as the persistence of symptoms following a minimum of two adequate trials of antipsychotics^2^. Clozapine is the only approved and evidence-based drug treatment for TRS, and is associated with higher adherence to treatment, decreased psychiatric hospital admission, reduced suicidal ideation and improvement of symptoms and outcomes^3-5^. Despite its clear benefits, clozapine is not always prescribed to those who could benefit from it. Retrospective analyses have shown that clozapine accounted for less than 5% of all antipsychotics prescribed for schizophrenia during 2006-2009 in the United States^6^, and that only a third of those eligible in the United Kingdom in 2019 are estimated to have received the drug^7^. Some important factors preventing wider clozapine use are a lack of clinician training and direct experience with clozapine, but most importantly concerns around its adverse drug reaction (ADR) profile^8^.

Health services in many countries mandate haematological monitoring for those receiving clozapine in order to prevent a rare and potentially fatal outcome, agranulocytosis, but there are many other potential ADRs that be challenging for clinical management^9^. Some of these, including constipation and seizures, can be life-threatening and are associated with clozapine metabolism, a complex interplay of biological processes that can also play a role in treatment outcomes^10^. Aiming to balance efficacy and ADR risk, guidelines recommend prescribing clozapine maintenance doses leading to plasma concentrations between 350-600 ng/mL, sometimes called a “therapeutic range”^11^. However, without regular assessments of clozapine pharmacokinetics (a framework known as “therapeutic drug monitoring”; TDM) the large inter-individual variability in clozapine metabolism makes the optimal prescription of safe and efficacious doses of clozapine a trial-and-error exercise^10^. Even robust predictors of variance in clozapine metabolism, for example smoking, co-medications, and weight, have small effects in practice, or are only useful for a subset of people and at certain stages of treatment^12^.

Pharmacogenomic approaches are established experimental designs for discovering novel predictors of drug metabolism and treatment response, often highlighting specific genetic markers among well-known biological pathways and enzymatic mechanisms^13^. Recent genome-wide association studies (GWAS) have pointed towards a small number of variants that could explain between 1% and 10% of the variance in clozapine pharmacokinetics after accounting for non-genetic factors^14,15^, and these may also have downstream relevance for ADRs^16^. Unfortunately, these studies have only been carried out in individuals of European ancestry, a clear limitation given the known diversity of drug-metabolising enzyme alleles worldwide, and the potential lack of transferability of genomic predictors across populations^13^. The Eurocentrism of clozapine pharmacogenomics studies is also particularly problematic as the prescription patterns and outcomes of this drug seem to be ancestrally stratified to some extent: For example, doses recommended for individuals of East Asian ancestry and indigenous populations of the Americas are lower than those typically prescribed to Europeans, to compensate for a generally slower clozapine metabolism in those populations^17^. Also, individuals of African ancestries may achieve greater therapeutic improvements than Europeans when prescribed clozapine, but are also more likely to discontinue the drug due to benign ethnic neutropenia, a condition likely caused by an African-specific and genetically-determined blood polymorphism^18^. Clearly, the importance of considering ancestral background in optimal clozapine prescribing in general, and pharmacogenomics research in particular, needs to be thoroughly investigated in large and ancestrally diverse samples. However, recruiting individuals on clozapine for genomic studies, either internationally or from populations that form minorities in Europe and North America, is a complex endeavour given the impairments and poor health caused by TRS, which act alongside a larger set of social and cultural barriers that influence participation and trust in clinical and academic schizophrenia research^19^.

Here, we seek to address ascertainment limitations in previous clozapine pharmacogenomics research and explore the ancestral diversity of common genetic variation relevant for clozapine metabolism. We report a cross-ancestry analysis of clozapine metabolism on 4,495 individuals linked both to genomic data and to 16,068 longitudinal blood monitoring assays. Using this dataset, we developed statistical models of clozapine pharmacokinetics and a conducted a GWAS on five ancestral biogeographic groups (Europeans, Sub-Saharan Africans, North Africans, Southwest Asians and East Asians), including admixed individuals. We then validated the use of our GWAS approach for deriving genomic predictors of clozapine metabolism and show for the first time that these are potentially transferrable across ancestral backgrounds.

## METHODS

### Samples

Pharmacokinetic and genomic data from the study participants were acquired as part of the CLOZUK study of individuals from the United Kingdom prescribed clozapine for the treatment of TRS. Specifically, the present study involves individuals present in the Zaponex® Treatment Access System (ZTAS), a clozapine monitoring service managed by the pharmaceutical company Leyden Delta (Nijmegen, Netherlands). Samples and data from these individuals were collected from ZTAS during the second (“CLOZUK2”; 2013-2015) and third waves (“CLOZUK3”; 2019-2021) of the study. Samples of European ancestry from CLOZUK2 have already been reported as part of genome-wide analyses of schizophrenia and clozapine metabolism^14^. CLOZUK3 and the non-European samples recruited from CLOZUK2 have also been reported in a study of clozapine prescription patterns^20^. The CLOZUK study received UK National Research Ethics Service approval (ref. 10/WSE02/15), following the UK Human Tissue Act.

### Selection of individuals for analysis

Curation of the CLOZUK pharmacokinetic and genomic data, including classification of individuals into biogeographic genetic ancestries, is detailed in the **Supplementary Note**. After these procedures, a total of 4,495 individuals (3268 males [72.7%] and 1227 females [27.3%]; mean age 42.19 years [range 18-85 years]) linked to 16,068 pharmacokinetic assays remained in the combined CLOZUK2 and CLOZUK3 dataset. A sample size breakdown by genetic ancestry and data collection wave is shown in **Table 1**. A detailed report on clozapine pharmacokinetics and doses for all genetic ancestry groups is available in **Supplementary Table 1**.

**Table 1.** Samples and clozapine pharmacokinetic assays included in the present study, stratified by data collection wave and genetic ancestry.

|  | *CLOZUK2* |  | *CLOZUK3* |  |
| --- | --- | --- | --- | --- |
| **Ancestry Group** | **No. of**<br>**Individuals** | **No. of**<br>**Assays** | **No. of**<br>**Individuals** | **No. of**<br>**Assays** |
| European | 2910 | 9213 | 767 | 3834 |
| Sub-Saharan<br>African | 192 | 598 | 51 | 242 |
| North African | 108 | 318 | 14 | 79 |
| Southwest Asian | 200 | 706 | 44 | 237 |
| East Asian | 36 | 112 | 5 | 18 |
| Admixed /<br>Unknown | 132 | 460 | 36 | 251 |
| **Total** | **3578** | **11407** | **917** | **4661** |

### Pharmacokinetic data analyses

Generalised linear mixed-effect model (GLMM) regression was used to assess and evaluate differences between ancestral groups in clozapine dosing and metabolism. Regression models were fitted with functions included in the *glmmTMB*^21^ and *ordinal*^*22*^ R packages, and appropriate probability distributions for each outcome were used as previously described^14^. These included the gamma distribution for plasma concentration outcomes (clozapine and norclozapine) and the normal distribution for the logarithms of the clozapine doses and the clozapine:norclozapine metabolic ratio. In all regressions, fixed effect covariates included the ancestry classification, the time between dose intake and blood sample (TDS), sex, age and age^2^. Additionally, clozapine dose was included as a fixed effect predictor in the clozapine, norclozapine and metabolic ratio regressions due to its strong linear correlation with these metrics^12^. One random effect predictor was fitted at the level of study participants, which allowed all the curated pharmacokinetic assays to be included into these regressions while preventing confounding due to repeated measurements (“pseudoreplication”)^23^. Due to their potentially heterogeneous genetic makeup, individuals classified as of “admixed/unknown” biogeographic ancestry were not used in these analyses.

### Genomic data analyses

GWAS of the clozapine and norclozapine plasma concentrations, and the metabolic ratio, were performed using TrajGWAS v0.13^24^, which uses a GLMM framework to model all individuals and longitudinal measurements in each analysis. This is an advance over previous approaches that required representative single phenotypes for each individual to be derived from the repeated measures^14,15,25^; as such constructs may not capture all the relevant variability of the data^26^. All TrajGWAS models included clozapine doses, TDS, sex, age and age^2^ as covariates. 10 genetic principal components (PCs) were included to control potential population stratification, as well as 4 genetic ancestry probabilities in place of the categorical ancestry variable as in previous cross-ancestry work^20^. The genome-wide significance level was set, as commonly defined in human genomics, at p≤5×10^−8^. Further details on fitting the TrajGWAS regression models and performing statistical fine-mapping to infer the number of causal variants in each locus (“k”) and the probability of each SNP of being a causal variant (“posterior probability of inclusion”, PPI) are given in the **Supplementary Note**.

### Polygenic scoring

To create independent training and testing sets for PRS analyses, the CLOZUK merged dataset was split in the CLOZUK2 and CLOZUK3 batches (**Table 1**). TrajGWAS was run in CLOZUK2 with identical models and parameters to the main analyses to generate training summary statistics. PRSice-2 v2.35 ^27^ was then applied to CLOZUK3 to generate clozapine, norclozapine and metabolic ratio PRS based on the CLOZUK2 results. To assess the effect of polygenicity in the PRS association analyses, 9 different SNP p-value inclusion thresholds were used to generate PRS: p<5×10^−8^, p<1×10^−5^, p<1×10^−4^, p<0.001, p<0.01, p<0.05, p<0.1, p<0.5 and p<1.

PRS regression was performed via GLMMs in R analogously to the pharmacokinetic analyses, using the full CLOZUK3 longitudinal dataset. For consistency with our GWAS approach, each GLMM assessing the effects of a single PRS incorporated as fixed-effect covariates the clozapine daily dose, TDS, sex, age, age^2^, 10 PCs and 4 genetic ancestry probabilities. A random-effect covariate was also introduced at the participant level. Further details on this procedure are given in the **Supplementary Note**.

## RESULTS

### Ancestral differences in clozapine pharmacokinetics

Significant differences (p<0.05) were observed for most of the pharmacokinetic comparisons between Europeans and the four non-European ancestry groups (**Table 2**). All non-European ancestries showed a slightly increased but significant clozapine:norclozapine metabolic ratio compared to Europeans, while the results for individual clozapine metabolites varied between ancestries.

**Table 2.** Ancestral differences in four outcome phenotypes related to clozapine pharmacokinetics as estimated with GLMM regression. Effect sizes (β) indicate whether individuals within each non-European ancestry have a greater (positive) or lower (negative) average value of the outcome than those of European ancestry, which are defined as the reference given their greater sample size (n=3677). Effect sizes of additional fixed-effect covariates of each model are given in **Supplementary Table 2**.

| Genetic Ancestry | Log(dose) |  |  | Clozapine |  |  | Norclozapine |  |  | Log(Metabolic Ratio) |  |  |
| --- | --- | --- | --- | --- | --- | --- | --- | --- | --- | --- | --- | --- |
| | $\beta$ | SE | p | $\beta$ | SE | p | $\beta$ | SE | p | $\beta$ | SE | p |
| Sub-Saharan African<br>(n=243) | -0.007 | 0.026 | 0.797 | -0.088 | 0.034 | 0.010 | -0.280 | 0.032 | $2.21 \times 10^{-18}$ | 0.191 | 0.017 | $1.73 \times 10^{-29}$ |
| North African (n=122) | -0.132 | 0.037 | $3.67 \times 10^{-4}$ | 0.045 | 0.048 | 0.346 | -0.025 | 0.045 | 0.585 | 0.072 | 0.024 | 0.003 |
| Southwest Asian (n=244) | -0.161 | 0.026 | $6.31 \times 10^{-10}$ | 0.135 | 0.034 | $5.91 \times 10^{-5}$ | 0.076 | 0.031 | 0.015 | 0.061 | 0.017 | $2.67 \times 10^{-4}$ |
| East Asian (n=41) | -0.162 | 0.063 | 0.010 | 0.162 | 0.082 | 0.049 | 0.055 | 0.077 | 0.473 | 0.113 | 0.041 | 0.006 |

GLMM effect sizes, controlled for dose and other factors, pointed towards slower clozapine metabolism in Asian individuals, as indexed by higher average clozapine plasma concentrations in those of East (β=0.162; SE=0.082; P=0.049) and Southwest Asian (β=0.135; SE=0.034; P=5.91×10^−5^) ancestries. Norclozapine plasma concentrations were also significantly higher in Southwest Asians (β=0.076; SE=0.031; P=0.015), an effect likely impacted by their higher clozapine levels that disappeared after controlling for clozapine plasma concentrations in this analysis (β=-0.008; SE=0.018; P=0.676). Consistent with these observations, in analyses of the clozapine doses prescribed throughout treatment, Asian individuals of both ancestries typically received lower doses of clozapine than Europeans (**Figure 1A**). These differences in plasma concentrations and doses had a similar magnitude to the effects of sex (**Supplementary Table 2**).

**Figure 1.**
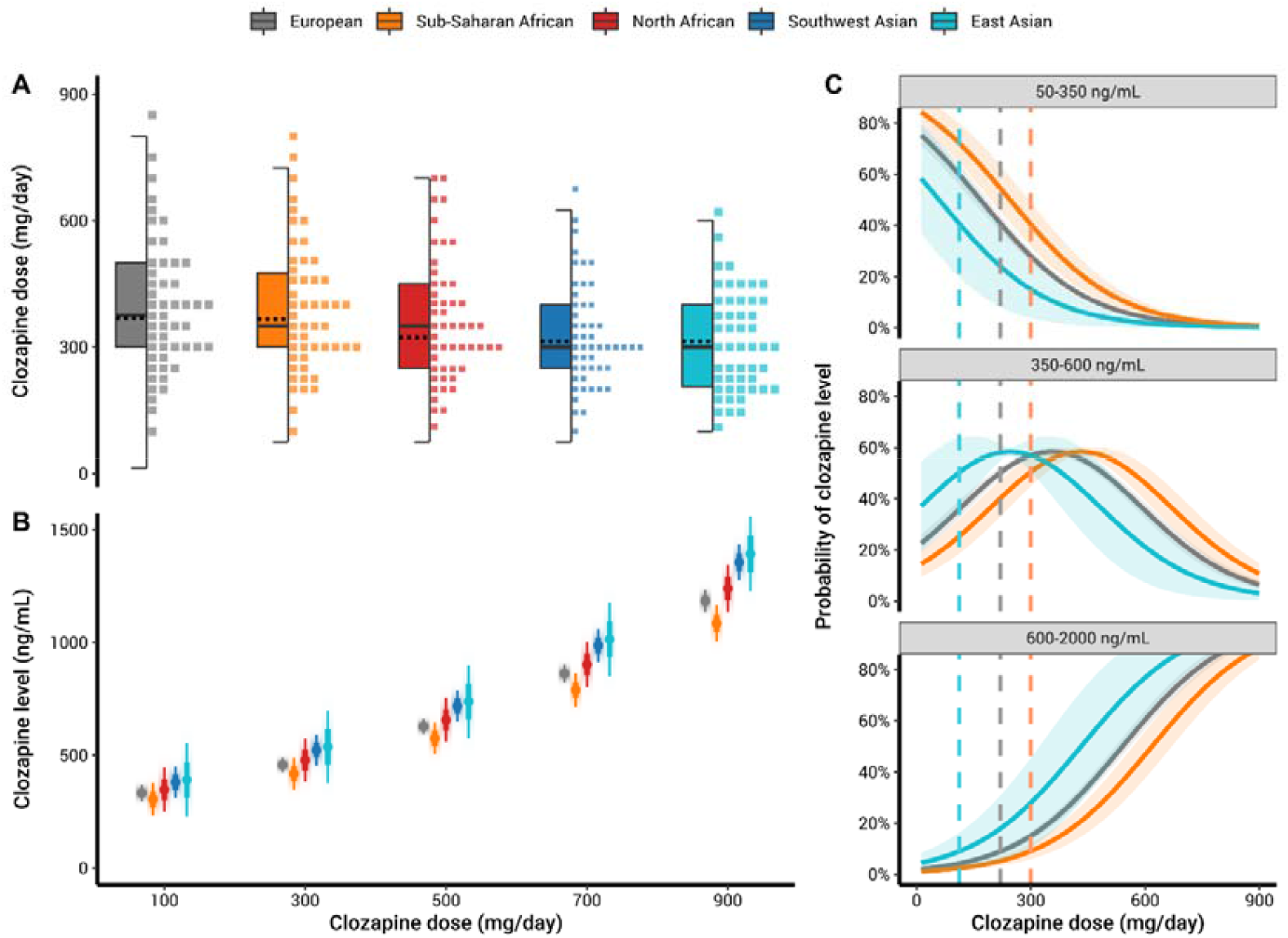
Analysis of clozapine pharmacokinetics in the CLOZUK longitudinal dataset. A: Distribution of clozapine doses throughout treatment, stratified by genetic ancestry; lines at the center of the boxplots indicate the median (continuous) or mean (dotted) of clozapine doses. B: Marginal effects of the ancestry groups in the relationship between clozapine doses and plasma concentrations (“levels”), indicated by a dot-whisker plot with mean levels at specific doses and 95% confidence intervals. C: Marginal effects of the ancestry groups in the relationship between clozapine doses and the probability of reaching clozapine levels inside or outside the therapeutic range (350-600 ng/mL).Shaded areas in the probability lines highlight a 95% confidence interval. Vertical dashed bars highlight the doses required by individuals in each ancestry group to reach the therapeutic range with 50% probability. For clarity, the North African and Southwest Asian groups have been omitted for this panel, a complete version can be found in **Supplementary Figure 6**.

Those of Sub-Saharan African ancestry showed the opposite results with regards to clozapine (β=-0.088; SE=0.034; P=0.010) and norclozapine plasma concentrations (β=-0.280; SE=0.032; P=2.21×10^−18^), which were lower relative to Europeans pointing towards faster metabolism (**Figure 1B**). There were no significant differences in average clozapine doses between those of Sub-Saharan African and European ancestries (β=-0.007; SE=0.026; P=0.797).

To indirectly assess whether these differences in prescriptions and metabolism might have implications for the effectiveness of clozapine, we used ordinal GLMMs to assess the probability of individuals within each ancestry having sub-therapeutic (<350 ng/mL), therapeutic (350-600 ng/mL) or supratherapeutic (>600 ng/mL) clozapine plasma concentrations throughout their treatment. Results of this model showed Southwest Asians to be approximately twice as likely to reach therapeutic or supra-therapeutic clozapine concentrations than Europeans throughout treatment (OR=1.974; SE=0.189; P=3.29×10^−4^), while Sub-Saharan Africans were half as likely to do so (OR=0.560; SE=0.193; P=2.67×10^−3^). Probability estimates of specific plasma concentration categories as a function of the clozapine dose also demonstrated the increased likelihood of sub-therapeutic concentrations among those of Sub-Saharan African ancestry in CLOZUK (**Figure 1C**). Specifically, these individuals required doses over 300 mg/day to reach the therapeutic interval with at least 50% probability (50.05%; SE=2.86%), while the same outcome was achieved at 220 mg/day in those of European ancestry (50.23%; SE=1.22%) or at 112 mg/day in East Asians (50.32%; SE=6.93%).

### Cross-ancestry SNPs and genes associated with clozapine metabolism

The results of the cross-ancestry GWAS of the plasma concentrations of clozapine and norclozapine, and of the clozapine:norclozapine metabolic ratio, can be seen in **Figure 2**. Across all phenotypes, 8 genome-wide significant (GWS) loci were found (**Table 3**), 5 of which have already been reported in Europeans^14,15^. We also noted novel cross-phenotype convergences in two of these known regions: *CYP1A1/1A2* (chromosome 15) and *UGT1A\** (chromosome 2). These loci were previously associated to clozapine and norclozapine respectively, but are associated to both phenotypes in our current analysis. We also found a novel signal for the metabolic ratio, indexed by the SNP rs41301394 (β= 0.196, SE= 0.035, p= 4.81×10^−8^), an intronic variant within *POR*, the gene encoding the NADPH:cytochrome P450 oxidoreductase protein. No additional genome-wide significant associations were seen in any of the ancestry-specific GWAS (**Supplementary Figures 1-4**), though the European-only analysis supported all our main results except for the locus tagging *POR*. The *CYP2C18* metabolic ratio association also surpassed the statistical threshold for genome-wide significance in the Southwest Asian-only analysis.

**Figure 2.**
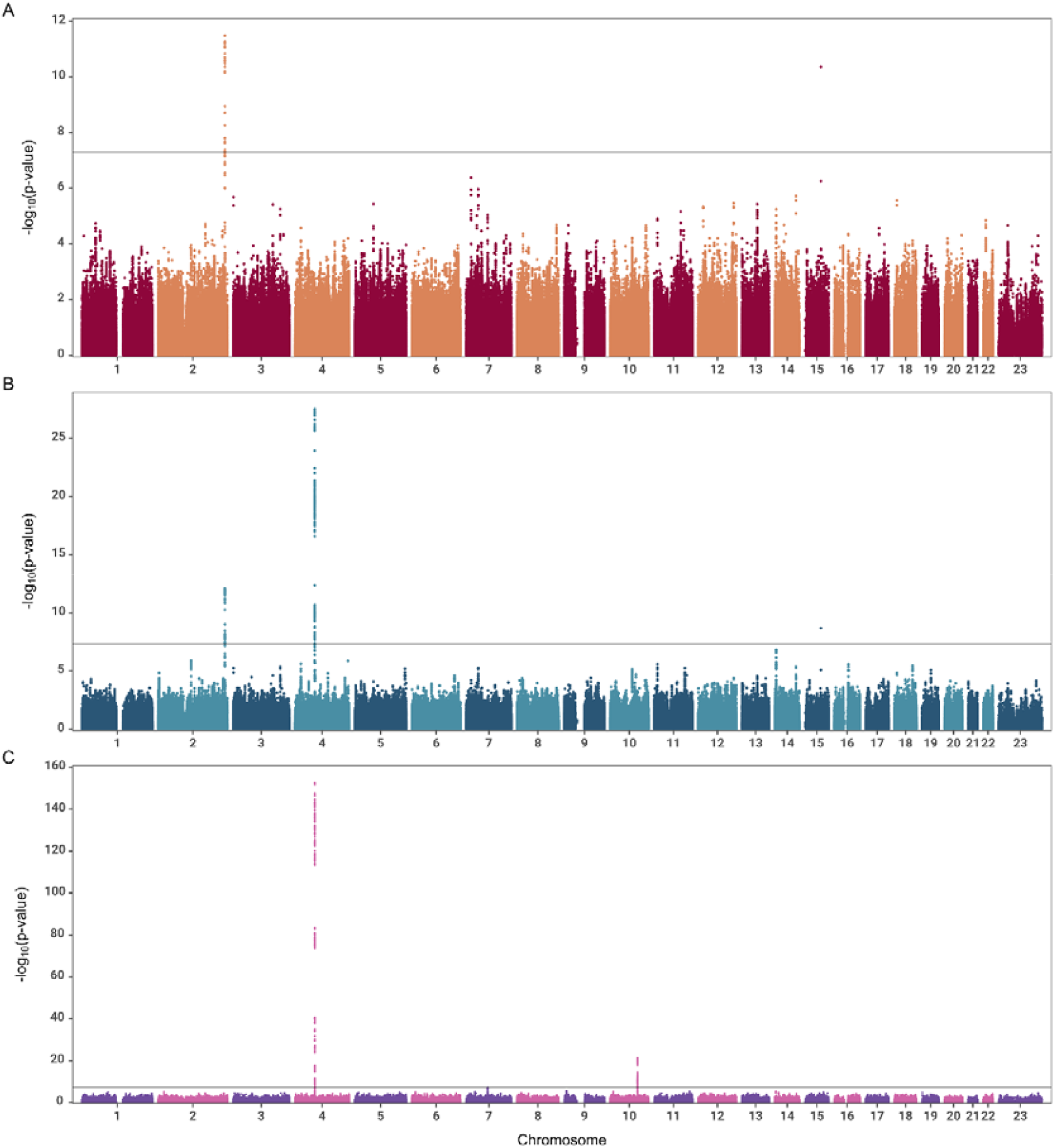
Manhattan plots of the GWAS analyses of clozapine metabolism carried out in the longitudinal CLOZUK dataset. Black horizontal line indicates the genome-wide significant p-value cutoff of 5×10^−8^. A: clozapine plasma concentrations (λ_GC_= 1.028). B: norclozapine plasma concentrations (λ_GC_= 1.018). C: clozapine:norclozapine metabolic ratio (λ_GC_= 1).

**Table 3.** Genome-wide significant loci associated with clozapine metabolism in the cross-ancestry CLOZUK sample using TrajGWAS. The closest gene to the index SNP (in GRCh37 coordinates) is highlighted in **bold**, with multiple genes highlighted for overlapping gene boundaries or in intergenic SNPs.

| Phenotype and Locus (containing all variants with $r^2 > 0.1$ to index) | Index SNP | p-value | Tagged genes |
| --- | --- | --- | --- |
| **Clozapine** |  |  |  |
| chr2:234610912-234676118 | rs3732218 | $3.26 \times 10^{-12}$ | *DNAJB3*, *MROH2A*,<br>*UGT1A1*, *UGT1A3*,<br>*UGT1A4*, ***UGT1A5***,<br>***UGT1A6***, ***UGT1A7***,<br>***UGT1A8***, ***UGT1A9***,<br>***UGT1A10*** |
| chr15:75019449-75027880 | rs2472297 | $4.39 \times 10^{-11}$ | ***CYP1A1***, ***CYP1A2*** |
| **Norclozapine** |  |  |  |
| chr4:69557365-70093260 | rs2926036 | $3.16 \times 10^{-28}$ | *UGT2A3*, *UGT2B4*,<br>*UGT2B7*, ***UGT2B10***,<br>*UGT2B11*, *UGT2B28* |
| chr2:234610912-234676118 | rs3732218 | $8.07 \times 10^{-13}$ | *DNAJB3*, *MROH2A*,<br>*UGT1A1*, *UGT1A3*,<br>*UGT1A4*, ***UGT1A5***,<br>***UGT1A6***, ***UGT1A7***,<br>***UGT1A8***, ***UGT1A9***,<br>***UGT1A10*** |
| chr15:75019449-75027880 | rs2472297 | $2.10 \times 10^{-9}$ | ***CYP1A1***, ***CYP1A2*** |

| **Metabolic ratio** |  |  |  |
| --- | --- | --- | --- |
| chr4:69603602-70366972 | rs835309 | 2.07x10<sup>-153</sup> | *UGT2A3, UGT2B4, UGT2B7, **UGT2B10**, UGT2B11, UGT2B28* |
| chr10:96024357-96848776 | rs1926711 | 4.74x10<sup>-22</sup> | *CYP2C8, CYP2C9, **CYP2C18**, CYP2C19, HELLS, NOC3L, PLCE1, TBC1D12* |
| chr7:75607155-75843524 | rs41301394 | 4.81x10<sup>-8</sup> | *MDH2, **POR**, SRRM3, STYXL1, TMEM120A* |

Aiming to identify credible causal SNPs and genes, we applied FINEMAP^28^ to each of the GWS loci. The best-fitting FINEMAP models for *CYP1A1/1A2* and *POR* included just a single causal variant (k=1); while *UGT1A*, UGT2B10* (in norclozapine) and *CYP2C18* had two (k=2). The metabolic ratio locus at *UGT2B10* could not be confidently fine-mapped with the default software settings (best-fitting k ≥ 5). We reran FINEMAP at this locus using a conditional analysis approach, which inferred a best causal model with k=7. Based on the data from the best FINEMAP model for each locus, we then defined credible sets with 95% posterior probability of including all causal SNPs at a locus. These contained between 1 and 411 variants, each with an estimated PPI value indicating its probability of being a causal variant (**Supplementary Table 3**). Apart from the metabolic ratio *CYP2C18* locus, all FINEMAP credible sets contained fewer SNPs than a previous fine-mapping effort^14^, though a more detailed comparison of fine-mapping results at the SNP level is provided in the **Supplementary Note**. Within credible sets, one SNP was confidently identified as the only causal signal behind the *CYP1A1/1A2* locus for both clozapine and norclozapine (rs2472297, PPI=1). SNPs with high causal probabilities (PPI ≥ 0.33) were also found within the boundaries of two genes: *UGT2B10* (norclozapine and metabolic ratio) and *POR* (metabolic ratio). These genes themselves cumulatively contained most of the PPI within each locus, suggesting they are the most likely genes in the region to be causally related to the association signal^29^.

To provide an estimate of the phenotypic effects of our discovered pharmacogenomic variants, we analysed the fine-mapped SNPs with the largest PPI at each phenotype/locus combination in R. Association statistics were very similar to the TrajGWAS results, with all markers retaining genome-wide significance (**Table 4**). Additionally, we also explored the cross-ancestry consistency of these markers by refitting the regression models within the CLOZUK ancestry groups. Across 40 tests (10 SNPs x 4 ancestries), we saw consistency (in direction and magnitude) of all SNP effect sizes with the main analysis in 38 instances, including every comparison in which the ancestry-specific association statistics were at least nominally significant (**Supplementary Table 4**).

**Table 4.** Effect sizes of fine-mapped genome-wide significant SNPs associated with clozapine metabolism phenotypes, in a ng/mL scale for plasma concentrations and a log scale for the ratio. PPI: Largest posterior probability of the variant being causal across FINEMAP and flashfm analyses. Unadjusted effect size estimates, controlled only for genomic covariates and a random effect, are given in **Supplementary Table 6**.

| SNP | Phenotype | Closest gene(s) | Effect allele | Other allele | Effect allele frequency | PPI | TrajGWAS | glmmTMB |  |  |
| --- | --- | --- | --- | --- | --- | --- | --- | --- | --- | --- |
| | | | | | | | p | $\beta$ | SE | p |
| **rs3732218** | Clozapine | *UGT1A5*<br>*UGT1A6*<br>*UGT1A7*<br>*UGT1A8*<br>*UGT1A9*<br>*UGT1A10* | A | G | 9.99% | 8.61% | $3.26 \times 10^{-12}$ | -0.125 | 0.0183 | $9.17 \times 10^{-12}$ |
| **rs2472297** | Clozapine | *CYP1A1*<br>*CYP1A2* | T | C | 23.91% | 100% | $4.39 \times 10^{-11}$ | -0.086 | 0.0132 | $7.60 \times 10^{-11}$ |
| **rs2926036** | Norclozapine | *UGT2B10* | G | A | 85.51% | 34.89% | $3.16 \times 10^{-28}$ | 0.165 | 0.0160 | $4.99 \times 10^{-25}$ |
| **rs115619871\*** | Norclozapine | *UGT2B10* | T | C | 9.82% | 6.71% | $2.50 \times 10^{-19}$ | -0.148 | 0.0173 | $1.26 \times 10^{-17}$ |
| **rs3732218** | Norclozapine | *UGT1A5*<br>*UGT1A6*<br>*UGT1A7*<br>*UGT1A8*<br>*UGT1A9*<br>*UGT1A10* | A | G | 9.99% | 10.77% | $9.36 \times 10^{-13}$ | -0.120 | 0.0172 | $2.66 \times 10^{-12}$ |
| **rs2472297** | Norclozapine | *CYP1A1*<br>*CYP1A2* | T | C | 23.91% | 100% | $2.10 \times 10^{-9}$ | -0.075 | 0.0124 | $1.04 \times 10^{-9}$ |
| **rs1902932** | Ratio | *UGT2B10* | A | G | 9.84% | 40.12% | $1.32 \times 10^{-123}$ | 0.171 | 0.0089 | $2.44 \times 10^{-83}$ |
| **rs115619871\*** | Ratio | *UGT2B10* | T | C | 9.82% | 36.88% | $2.30 \times 10^{-123}$ | 0.171 | 0.0089 | $4.33 \times 10^{-83}$ |
| **rs76413136** | Ratio | *CYP2C18* | T | C | 23.25% | 10.60% | $9.18 \times 10^{-22}$ | 0.060 | 0.0065 | $2.75 \times 10^{-20}$ |
| **rs41301394** | Ratio | *POR* | T | C | 26.79% | 43.68% | $4.81 \times 10^{-8}$ | -0.034 | 0.0061 | $2.27 \times 10^{-8}$ |

\* Putatively shared causal variant for norclozapine and the clozapine:norclozapine metabolic ratio reported by flashfm.

### Genomic prediction of clozapine metabolite concentrations in independent datasets

All clozapine metabolism PRS generated from CLOZUK2 were associated, at several p-value thresholds, with their respective phenotypes in CLOZUK3, explaining a maximum of 0.61% (clozapine), 1.59% (norclozapine) and 7.26% (metabolic ratio) of the variance after accounting for fixed-effect and random-effect predictors (**Supplementary Table 5**). For all phenotypes, PRSs displaying the stronger association and greater variance explained were those built only with GWS SNPs, and all associations at this p-value threshold remained significant after splitting the testing dataset by European/non-European ancestry (**Supplementary Table 5**). We also compared this PRS predictor with one analogously generated from our 2019 European-only GWAS^14^. Interestingly, the cross-ancestry PRS predictor showed improved performance even beyond the GWS threshold, with all significant associations from the 2019 PRS being replicated and strengthened in our current study, which also included novel significant results (**Figure 3**). Many of these associations also replicated with similar effect sizes in the two largest non-European ancestries within CLOZUK3, the Sub-Saharan Africans and the Southwest Asians, supporting the transferability of our PRS as predictors of clozapine metabolism to underrepresented populations in genomic research (**Supplementary Figure 5**).

**Figure 3.**
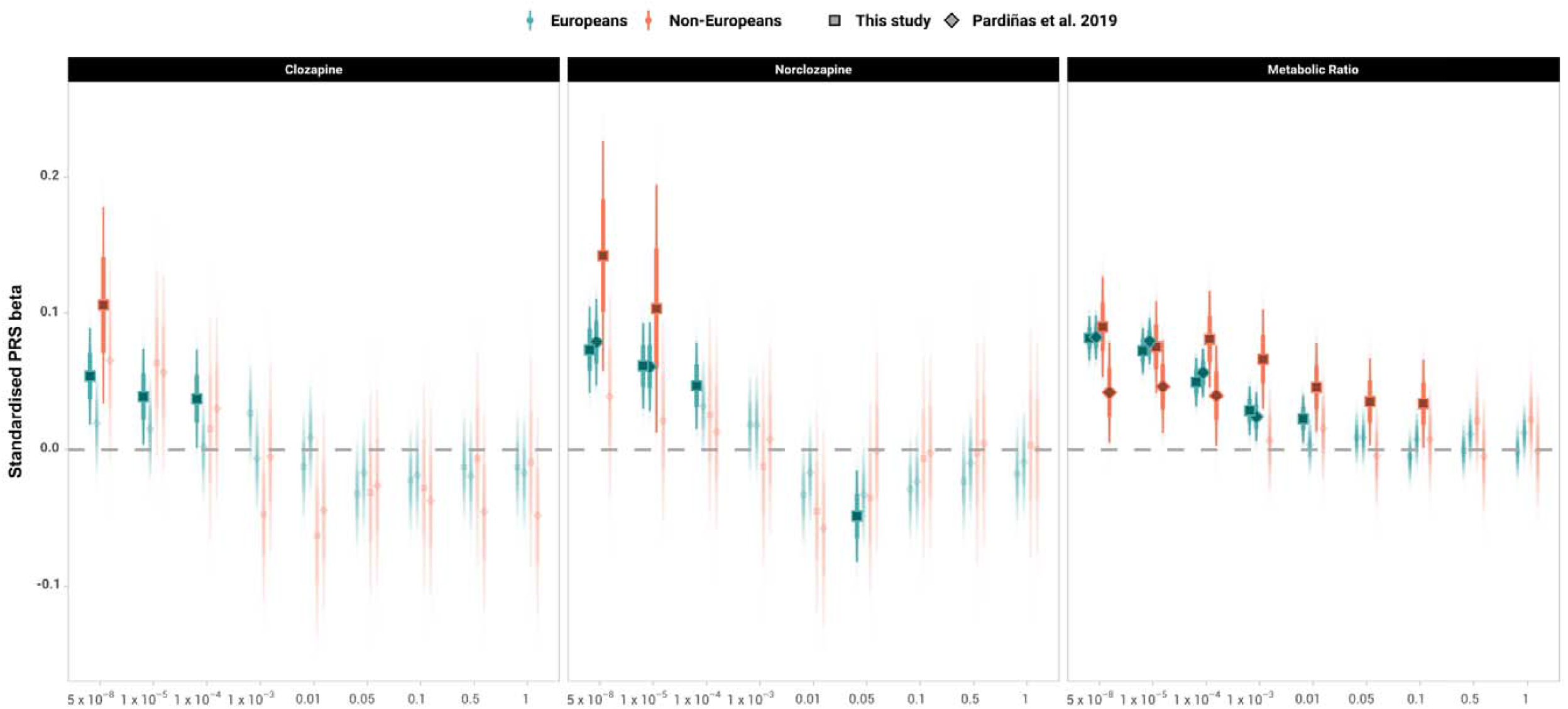
Association between polygenic scores (PRS) for clozapine metabolism generated from a CLOZUK2 GWAS and their corresponding phenotypes in the CLOZUK3 longitudinal dataset. Dots and whiskers indicate the estimated value of the regression effect size and its corresponding 95% confidence interval. Bold colours indicate statistically significant associations; semi-transparent colours indicate non-significant effect sizes.

## DISCUSSION

This study is, to our knowledge, the first to explore clozapine metabolism and pharmacogenomics using a diverse cross-ancestry sample and statistical methods that fully take advantage of the longitudinal nature of clozapine monitoring assays. We show using pharmacokinetic data that, on average, those of Sub-Saharan African ancestry were more likely than those of European ancestry to be fast clozapine metabolisers but were not prescribed different doses to those of European ancestry (**Table 2, Figure 1**). Further analysis also showed that those of Sub-Saharan African ancestry were the least likely in our dataset to achieve therapeutic plasma concentrations at clozapine doses below 300 mg/day, which parallels recent findings in non-clozapine antipsychotics^30^. These results were unexpected and of potential clinical importance given that the most recent revision of clozapine treatment guidelines does not report pharmacokinetic differences between people of European and African or African-American ancestry^17^, though the latter display increased rates of clozapine discontinuation and lower rates of treatment adherence in comparison to other ethnicities and ancestries^19^. Given the evidence supporting that the clozapine “therapeutic range” is consistently and cross-ancestrally associated with treatment response^11^, sub-optimal prescriptions to potential fast metabolisers are likely to lead to modest treatment response and unnecessary exposure to ADRs. This supports the argument that robust assessments of clozapine metabolism, including TDM, should be carried out across ancestries and populations to inform accurate and safer dosing practices for the diverse real-world pool of potential clozapine users^17^.

Our longitudinal cross-ancestry GWAS approach provides evidence that loci previously identified in Europeans also have pharmacokinetic associations in other ancestral backgrounds, a reassuring finding as some of these variants are only found at low frequencies in most of the world’s populations and thus are likely underpowered for cross-ancestry testing (**Supplementary Table 3, Supplementary Table 4**). In particular, the evidence for association between each of rs3732218 (*UGT1A\**) and rs2472297 (*CYP1A1/1A2*) with both clozapine and norclozapine plasma concentrations, reinforces that these represent or at least index causal variants influencing pharmacokinetics (**Table 3**). The fact that the directions of effect of both variants are congruent across all analyses suggests primary effects on clozapine pharmacokinetics might be mediating SNP effects on norclozapine levels; but statistically evaluating and quantifying this is not straightforward in a GLMM^31^. In any case, the association of these variants places a putative mechanism of action for clozapine pharmacogenomics early in the clozapine metabolism pathway, perhaps at the point where serum free clozapine is first demethylated or glucuronidated in the liver. Multiple and complex biochemical processes could be at play, as the *UGT1A\** family (particularly *UGT1A4*) has been linked to the excretion of both norclozapine and clozapine^32^; and *CYP1A2* drives the clozapine to norclozapine conversion and participates in producing clozapine N-oxide^33^, a secondary metabolite that is primarily formed at high concentrations of the drug. Thus, gaining further insight into these associations requires direct experimental validation.

A new genome-wide significant locus for the clozapine:norclozapine metabolic ratio was identified in *POR* indexed by rs41301394, an intronic SNP with high posterior probability of being the causal variant within this locus (PPI=0.437). This is already a known pharmacogenomic marker that is associated with variation in warfarin maintenance doses in East Asians^34^, although with only preliminary (“class 3”) levels of evidence as annotated by the PharmGKB database. The gene itself is, however, robustly implicated in xenobiotic metabolism, is ubiquitously expressed in human tissues, and is an essential part of all CYP monooxygenase protein complexes^35^. Given the lack of effect of rs2472297 (*CYP1A1/1A2*) in the clozapine:norclozapine ratio GWAS (β= - 0.039, SE= 0.022, p= 0.084), this finding suggests that the metabolic ratio might be more strongly related to the pharmacokinetics of norclozapine (with which it shares a common signal at *UGT2B10*) than to clozapine plasma concentrations. This supports a recent review pointing out that the metabolic ratio does not relate to direct metrics of clozapine metabolism^36^, and that its use in TDM frameworks and research studies as an index of CYP1A2 activity should be re-evaluated.

Finally, our polygenic analyses demonstrate the utility of this methodology to generate pharmacogenomic predictors from GWAS of drug metabolism. This is despite these traits likely having an oligogenic genetic architecture (**Supplementary Note**), as seen in many other metabolic traits, instead of the polygenic basis often exploited in most PRS-based research. In an oligogenic framework, phenotypic prediction using PRS is expected to work best at conservative SNP p-value thresholds as we observe through our tests^37^, with the paucity of variants considered in these metrics being offset by relatively large per-allele effect sizes (e.g. one minor allele of rs2472297 has an effect on clozapine plasma concentrations roughly equivalent to reducing the dose in 50 mg/day^14^). Indeed, we show that all of our clozapine metabolism PRS were associated cross-ancestrally with their respective phenotypes in independent samples, and that their variances explained, albeit small, were consistent with those previously estimated for index SNPs within GWS loci^14^. A direct comparison with our previous study supports that our novel longitudinal GWAS methodology and more diverse samples contribute to increasing the power of this approach particularly in those of non-European ancestries (**Figure 3, Supplementary Table 5, Supplementary Figure 5**), leading to a PRS predictor potentially transferrable across ancestries. This is particularly promising for future prospective trials as only small panels of SNPs might be needed to evaluate the relevance of these genomic findings for clinical and prescribing interventions, facilitating the design of these studies and the targeting of large and global samples.

While this study benefited from data from a large-scale genetic study on TRS, the fact that CLOZUK samples were solely recruited within the UK meant that only relatively limited sample sizes were available for individuals of non-European ancestries. We have attempted to maximise the use of this data by only making ancestry-based exclusions when needed to avoid potential confounding, and by incorporating as much data as possible into pharmacokinetic and genomic analyses using GLMM regression. However, it is likely that the GWAS we carried out in non-European ancestries were still too underpowered to detect ancestry-specific associations at the genome-wide significant threshold, although they showed a notable consistency at the level of direction of effects for the main reported signals (**Supplementary Table 4**). Finally, the lack of information in CLOZUK on known predictors of clozapine metabolism, most importantly smoking habits, weight, and concomitant medication; is another limitation of the study. However, the main reported effect of not accounting for these exposures in genomic studies of metabolic traits, including clozapine, is the masking of signals^15,38^. This is arguably a lesser concern than potential false positive findings but still one which should be considered when evaluating our work in a broader context, as it could for example downsize the utility or variance explained by SNPs or PRS.

In summary, this study adds to the evidence behind previously discovered associations related to clozapine pharmacokinetics and establishes cross-ancestral convergences in pharmacogenomic markers for the first time in clozapine research. It also identifies that the slower clozapine metabolism previously reported for individuals of East Asian ancestry is likely to also apply to Southwest Asians, while fast clozapine metabolism appears common in Sub-Saharan Africans. It follows that clozapine dosing and titration protocols developed using data from populations of European descent are unlikely to be optimal for a substantial proportion of humanity, and current clinical practice should be assisted by TDM approaches whenever possible. Indeed, studies conducted in the US highlight that clozapine underutilization is currently greatest in all non-White groups^19^. Our results therefore contribute to the knowledge gap in the identification of robust predictors of clozapine metabolism that could ultimately be used to design interventions seeking to improve the access to this drug, and its safety, for everyone eligible. While our novel findings and interpretations require replication, our study in overall shows a clear benefit of using indexes of genetic ancestry as a necessary part of pharmacological research. As the pharmacogenomics community strives to incorporate diverse populations into its routine work^39^, the future shows promise for the development of personalised medicine initiatives for clozapine treatment, which could ensure that this drug brings safe benefits to a larger and more diverse population of individuals with TRS.

## Data Availability

To comply with the ethical and regulatory framework of the CLOZUK project, access to individual-level data requires a collaboration agreement with Cardiff University. Requests to access these datasets should be directed to Prof. James T. R. Walters.

## ACKNOWLEDGEMENTS

AFP and DBK were supported by an Academy of Medical Sciences “Springboard” award (SBF005\1083). The CLOZUK study was supported by the following grants from the Medical Research Council to Cardiff University: Centre (MR/L010305/1), Program (MR/P005748/1), and Project (MR/L011794/1, MC_PC_17212); as well as the European Union’s (EU) Seventh Framework program (279227, “CRESTAR”). This work acknowledges the support of the Supercomputing Wales project, which is partly funded by the European Regional Development Fund (ERDF) via Welsh Government. It also acknowledges Lesley Bates, Catherine Bresner and Lucinda Hopkins (Cardiff University) for laboratory sample management, as well as Andy Walker (Magna Laboratories), Anoushka Colson (Leyden Delta) and Hreinn Stefansson (deCODE Genetics) for contributing to the sample collection, anonymization, data preparation and genotyping efforts of the CLOZUK2 sample.

## CONTRIBUTORS STATEMENT

AFP, DBK and JW conceived and designed the study. AK, JJ, and MH contributed samples for genotyping. AK contributed and verified pharmacokinetic data. AFP, DBK, MR and FT and LMSJ curated genotypic data and performed statistical and genomic analyses. AFP had access to all data and supervised its overall curation and analysis. AFP, DBK, MR, MO, MCOD, and JTRW participated in the primary drafting of the manuscript. All authors had the opportunity to review and comment on the manuscript, and all approved the final manuscript.

