## Supplementary Note for "Pharmacokinetics and pharmacogenomics of clozapine in an ancestrally diverse sample: A longitudinal analysis and GWAS using clinical monitoring data from the UK"

#### Supplementary Methods

##### Curation of the pharmacokinetic data

19,096 pharmacokinetic assays were available for 4,760 CLOZUK individuals, including information on the clozapine and norclozapine plasma concentrations, the daily clozapine dose, and the time of both the drug intake and blood draw. All plasma concentrations were determined by a standard high-performance liquid chromatography mass spectrometry (HPLC-MS) procedure at Magna Laboratories (Ross-on-Wye, U.K.), and further details are provided in a previous publication (Pardiñas et al. 2019). The pharmacokinetic assay dataset was assessed for potential clerical errors (e.g. negative time periods, multiple samples recorded at the same time point) and cross-checked with ZTAS records to ensure data integrity (e.g. correct year of birth for each individual). Data from all assays containing any erroneous information was removed. Additionally, assays were excluded from further analyses if they fit any of the following criteria: (i) blood drawn outside of a "trough sample" interval of 6- to 24-hours postdose (Flanagan 2010); (ii) clozapine or norclozapine concentration <50 ng/mL, outside the minimum detection range of the HPLC-MS instrument; (iii) clozapine concentration >2000 ng/mL, reaching the range of potential toxicity (Flanagan et al. 2020); (iv) clozapine:norclozapine ratio outside of the 0.5-3.0 interval, indicating potential non-adherence to treatment (Ellison and Dufresne 2015); and (v) clozapine dose >900 mg/day, the maximum advised by the British National Formulary. Finally, due to potentially different treatment regimes, we also removed all data from individuals under 18 years of age.

##### Curation of the genomic data

The CLOZUK2 samples underwent genotyping, quality control and imputation as previously described (Pardiñas et al. 2019). Briefly, 7,417 samples were genotyped by deCODE Genetics (Reykjavík, Iceland), using an Illumina HumanOmniExpress-12 array with 719,665 SNPs. Quality control was performed using PLINK v1.9 (Chang et al. 2015). Samples and markers with a missingness rate of >2% and samples with an inbreeding coefficient of F>0.2 were excluded from further analyses. After curation and merging with the pharmacokinetic assay dataset, 3,578 samples genotyped at 698,442 SNPs remained in CLOZUK2.

For CLOZUK3, 1,439 samples were genotyped at the Icahn School of Medicine at Mount Sinai (New York City, USA) using an Illumina® Infinium Global Screening Array-24 (GSA-24) with 654,027 SNPs. The curation of both samples and markers was performed using the same procedures as CLOZUK2, implemented in the DRAGON-Data pipeline (Hubbard et al. 2022).

After curation and merging with the pharmacokinetic assay dataset, 917 samples genotyped at 537,334 SNPs remained in CLOZUK3.

Genotype imputation for both cohorts was performed using the Haplotype Reference Consortium (HRC) panel through the Michigan Imputation Server (McCarthy et al. 2016). All imputed genotype dosages of CLOZUK2 and CLOZUK3 were further curated within each cohort with the following parameters: Imputation quality $r^2 \geq 0.7$; hard-call genotype probability ≥ 80%; hard-call missingness ≤ 5%; minor allele count (MAC) ≥ 2. The remaining variants in common between both samples were then merged and a second round of dosage curation was performed with the following parameters: Hard-call missingness ≤ 2%; MAC ≥ 400 and Hardy-Weinberg equilibrium (HWE) mid-p > $10^{-6}$. This led to over 2.9 million high-quality markers for GWAS and PRS analyses. For context, the choice of setting quality control thresholds on MAC instead of minor allele frequency (MAF) was motivated by evidence relating this parameter to the power and robustness of multiple genomic association tests (Ma et al. 2013; Ray and Chatterjee 2020), for which usual MAF thresholds are not always relevant.

**Prediction of biogeographic genomic ancestry**

Genomic data was also used for biogeographic ancestry prediction using a linear discriminant analysis (LDA) model based on ancestry-informative markers (AIMs; Legge et al. 2019). Briefly, genotype data was merged with a public geo-localised reference panel based on the Human Genome Diversity Project (HGDP; Li et al. 2008). The population differentiation statistic $F_{ST}$ was used to identify AIMs by selecting those overlapping SNPs maximally differentiated between HGDP superpopulations. Genotype principal components (PCs) were then generated based on these AIMs and processed through a LDA model with the HGDP samples of known ancestry as training set and CLOZUK2/CLOZUK3 as test sets. The model resulted in a set of probabilities, for each sample, of belonging to each of five global biogeographic ancestries: “European”, “North African”, “Sub-Saharan African”, “East Asian” and “Southwest Asian” (Legge et al. 2019). For comparability with other studies, it should be noted that these ancestries are equivalent in definition to the non-admixed standardised biogeographic groups recently proposed for use in pharmacogenomics research (Huddart et al. 2019). Individuals were assigned to one specific ancestry if their LDA probability surpassed a threshold of 80%, and to a group labelled as “admixed/unknown” otherwise. For carrying out ancestry-specific GWAS, given the small sample size of some of these subgroups, the merged

CLOZUK dosages were curated within each ancestry with the following parameters: Hard-call missingness ≤ 2%; hard-call MAC ≥ 40 and HWE mid-p > $10^{-4}$.

**GWAS model fitting and statistical fine-mapping**

For our main GWAS analyses, we estimated effect sizes, standard errors and p-values using the Wald test implemented in TrajGWAS (Ko et al. 2022). This uses the "within-subject variance estimator by robust regression" (WiSER) approach (German et al. 2021), a GLMM framework that can accommodate predictors of the outcome mean and variance while being robust to multiple outcome and random effect distributions. However, computational problems with the WiSER procedure may occur while modelling long-tailed outcome distributions, requiring changing the numerical optimisation procedures for particular genomic regions or SNPs (Ko et al. 2022). As long tails and extreme values are often features of pharmacokinetic metrics (Lindsey, Jones, and Jarvis 2001), we sought to avoid this potential problem by normalising and standardising all of our outcomes before the TrajGWAS analysis. For this we used a cube root transformation for the gamma-distributed clozapine and norclozapine concentrations (Wilson and Hilferty 1931; Terrell 2003), and a logarithmic transformation for the log-normally-distributed metabolic ratio. All covariates were also standardised, as recommended to improve the computational performance of GLMMs (Gelman, Hill, and Vehtari 2020). Both procedures were also used in the genomic *glmmTMB* analyses.

Taking advantage of the WiSER modelling capabilities, TrajGWAS models included sex as a predictor of both the between-person mean and the within-person variance, reflecting the observation that differential smoking habits in males and females might account for some of the "noise" in clozapine pharmacokinetic assays (Diaz et al. 2005). When replicating the results of specific SNPs in *glmmTMB* (**Table 4**), we approximated this approach by setting sex as a predictor of both the mean ("location") and the residual variance ("scale") in the GLMM.

Ancestry-specific GWAS were carried out using the saddlepoint approximation (SPA) to the score test also implemented in the TrajGWAS, which is appropriate even for small sample sizes and rare allele frequencies (Ko et al. 2022). These analyses did not include ancestry probabilities as covariates, though still included the full set of 10 PCs to control for population stratification. Ancestry-specific GWAS were not run in the sample of individuals classified as "admixed/unknown" due to their potential ancestral complexity; nor in those of East Asian ancestry due to the small sample size of this group (n=41).

Genomic inflation statistics (λ values) were calculated on the output of each GWAS using the *fastman* R package (Paria, Rahman, and Adhikari 2022). LD clumping was also performed to summarise the results and identify tagged genes. LD clumps were formed around genome-wide significant ("index") SNPs and variants with $r^2>0.1$ within 3000 kb were assigned to these clumps. Fine-mapping of genome-wide significant clumps was carried out with FINEMAP v1.41 (Benner et al. 2016) using a maximum number of k=5 causal SNPs and locus-wide LD panels created from the CLOZUK dosage data. Clumps in common between different outcomes were additionally run through flashfm v1.0 (Hernández et al. 2021) using the top 1,000 causal configurations identified by FINEMAP, in order to pinpoint potentially shared causal SNPs. Additionally, for each genome-wide significant GWAS locus and phenotype, dosages from the fine-mapped SNP with the largest probability of being causal were extracted and analysed in R (using *glmmTMB*) to estimate GLMM effect sizes in the scale of the original pharmacokinetic variables.

**Estimation of SNP-based heritability**

We used MiXeR v1.3 to estimate the heritability of clozapine metabolism phenotypes using summary statistics, given its reported good performance across a range of genomic architectures including oligogenic traits (Frei et al. 2019). An LD reference panel was generated directly from CLOZUK best-guess genotypes and all analyses were ran using default parameters. As recommended by the software developers, results were generated from 20 MiXeR runs using random sets of ~500,000 LD-independent SNPs (MAF > 0.05; $r^2 < 0.9$).

**Polygenic score association analysis**

As described in the main text, GLMM regression models to estimate PRS effect sizes followed the main pharmacokinetic analyses. To account for multiplicity of tests throughout this procedure, PRS association p-values were corrected using false discovery rates (FDR) within each outcome, and considered significant at a threshold of FDR $p \leq 0.1$ (Benjamini and Hochberg 1995). As an index of the proportion of variance explained by PRS in the context of other fixed and random effects, a semi-partial $R^2$ statistic was estimated in R using *partR2* (Stoffel, Nakagawa, and Schielzeth 2021). Given the current implementation of this method cannot accommodate gamma-distributed GLMMs, all PRS models were re-fitted for $R^2$ estimation using normalised phenotypes. Following a recent recommendation by Rights and Sterba (2020), time-varying fixed-effect predictors (age, $age^2$, clozapine daily dose and TDS) were also centred at the individual level ("de-meaned") in the re-fitted LMM models.

### Supplementary Results

#### Support for SNPs previously associated to clozapine metabolism through GWAS

Several variants have been highlighted before as putative pharmacogenomic markers of clozapine metabolism in European ancestry studies (Pardiñas et al. 2019; Smith et al. 2020). These include a SNP intergenic to *CYP1A1/CYP1A2* (rs2472297), fine-mapped missense variants in *UGT2B10* (rs61750900), *UGT1A4* (rs2011425) and *CYP2C18* (rs1126545), and an intronic eQTL in *NFIB* (rs28379954). From this list, our credible sets defined by FINEMAP and flashfm included rs2472297 (largest PPI=1), rs2011425 (largest PPI=0.026) and rs1126545 (largest PPI=0.021). The other SNPs were not found within these results. On the *UGT1A4* locus, we instead found several other missense SNPs within the FINEMAP credible sets, but the majority were within a *UGT1A5* exon (4/8 for clozapine, cumulative PPI=0.119; 4/7 for norclozapine, cumulative PPI=0.110). This result was consistent with the smaller flashfm credible sets, which had most of their missense variants also mapped to *UGT1A5* (5/6 for clozapine, cumulative PPI=0.127; 5/6 for norclozapine, cumulative PPI=0.120). On the *UGT2B10* locus, for either norclozapine or the metabolic ratio, none of our analyses included any missense variants as part of credible sets, and in fact those were largely different between the assessed phenotypes. It should be noted this result could have been confounded by the large number of causal SNPs inferred for the metabolic ratio GWAS, which itself might have arisen due to the complexity and breadth of the association signal (1385 SNPs). Finally, rs28379954 could not be evaluated in our GWAS due to the poor imputation quality of the *NFIB* gene within CLOZUK2 and CLOZUK3 (imputation $r^2 < 0.6$).

Additional analyses using flashfm were undertaken for loci shared between multiple phenotypes. This led to further shrinking in the *UGT1A** credible set, from 90-107 SNPs in the norclozapine and clozapine analyses, respectively, to 74 SNPs (**Supplementary Table 3**). However, the credible sets of *CYP1A2* and *UGT2B10* widened, likely because flashfm uses information from all the FINEMAP models, not just those that are the best-fit, and thus reflects uncertainty in the number of causal variants underpinning the GWAS signal. Using flashfm also revealed a set of SNPs in high LD ($r^2$>0.7) within the *UGT2B10* locus that were putatively causal for both phenotypes and partially within the coding region of this gene (SNP group "A"; norclozapine cumulative PPI=0.169; ratio cumulative PPI=1; **Supplementary Table 3**).

**Heritability and polygenicity of clozapine metabolism**

The univariate analyses models implemented in MiXeR supported the oligogenicity of clozapine metabolism, with less than 0.001% of genetic variants genome-wide having non-zero effects for any of the analysed phenotypes. Common SNP-based heritability values were estimated at 2.87% (SE=0.30%) for clozapine; 4.86% (SE=0.28%) for norclozapine and 8.62% (SE=0.59%) for the metabolic ratio. Despite these low values, all the AIC/BIC values for these models were positive with the only exception of the BIC value of clozapine, supporting that the current TrajGWAS analysis is sufficiently powered for this procedure.

#### Supplementary Table 2

Additional fixed-effect covariates for the four clozapine pharmacokinetics models reported in **Table 2**, as estimated with GLMM regression. Predictor names indicate their unit of measurement or, in the case of binary predictors, their non-reference level. Effect sizes (β) indicate the positive or negative impact of a one-unit increase of the predictor in the average of the outcome, across all the individuals and longitudinal assays of the cross-ancestry CLOZUK sample.

| **Predictor** | **Log(dose)** | | | **Clozapine** | | | **Norclozapine** | | | **Log(Metabolic Ratio)** | | |
|---|---|---|---|---|---|---|---|---|---|---|---|---|
| | **β** | **SE** | **p** | **β** | **SE** | **p** | **β** | **SE** | **p** | **β** | **SE** | **p** |
| Daily dose (mg/day) | - | - | - | 0.002 | $3.60 \times 10^{-5}$ | $4.02 \times 10^{-426}$ | $6.01 \times 10^{-4}$ | $2.13 \times 10^{-5}$ | $5.26 \times 10^{-175}$ | $-8.12 \times 10^{-5}$ | $1.82 \times 10^{-5}$ | $8.01 \times 10^{-6}$ |
| Time between dose and blood draw (hours) | -0.003 | 0.0010 | 0.009 | -0.010 | 0.0015 | $4.77 \times 10^{-10}$ | 0.005 | 0.0009 | $7.64 \times 10^{-9}$ | -0.010 | 0.0008 | $2.94 \times 10^{-34}$ |
| Sex (male) | 0.129 | 0.0137 | $8.35 \times 10^{-21}$ | -0.147 | 0.0179 | $1.58 \times 10^{-16}$ | -0.032 | 0.0096 | $7.87 \times 10^{-4}$ | -0.015 | 0.0089 | 0.102 |
| Age (years) | -0.001 | 0.0005 | 0.051 | 0.004 | 0.0007 | $1.34 \times 10^{-9}$ | $9.77 \times 10^{-4}$ | 0.0004 | 0.010 | $7.09 \times 10^{-4}$ | 0.0004 | 0.043 |
| $Age^2$ ($years^2$) | $-2.93 \times 10^{-4}$ | $3.45 \times 10^{-5}$ | $2.14 \times 10^{-17}$ | $6.61 \times 10^{-6}$ | $4.60 \times 10^{-5}$ | 0.886 | $4.24 \times 10^{-5}$ | $2.50 \times 10^{-5}$ | 0.090 | $-3.22 \times 10^{-5}$ | $2.30 \times 10^{-5}$ | 0.161 |
| Batch (CLOZUK3) | -0.060 | 0.0148 | $4.78 \times 10^{-5}$ | 0.022 | 0.0190 | 0.236 | 0.032 | 0.0102 | 0.002 | -0.017 | 0.0094 | 0.065 |

#### Supplementary Table 6

Effect sizes of fine-mapped genome-wide significant SNPs associated with clozapine metabolism phenotypes, in a ng/mL scale for plasma concentrations and a log scale for the ratio. Only genomic covariates (principal components and ancestry probabilities) and an individual-level random effect have been included in these models. PPI: Largest posterior probability of the variant being causal across FINEMAP and flashfm.

| SNP | Phenotype | Closest gene(s) | Effect allele | Other allele | Effect allele frequency | PPI | TrajGWAS | glmmTMB | | |
|---|---|---|---|---|---|---|---|---|---|---|
| | | | | | | | p | β | SE | p |
| **rs3732218** | Clozapine | *UGT1A5 UGT1A6 UGT1A7 UGT1A8 UGT1A9 UGT1A10* | A | G | 9.99% | 8.61% | $3.26 \times 10^{-12}$ | -0.085 | 0.0183 | $3.17 \times 10^{-6}$ |
| **rs2472297** | Clozapine | *CYP1A1 CYP1A2* | T | C | 23.91% | 100% | $4.39 \times 10^{-11}$ | -0.066 | 0.0132 | $5.52 \times 10^{-7}$ |
| **rs2926036** | Norclozapine | *UGT2B10* | G | A | 85.51% | 34.89% | $3.16 \times 10^{-28}$ | 0.159 | 0.0165 | $6.61 \times 10^{-22}$ |
| **rs115619871*** | Norclozapine | *UGT2B10* | T | C | 9.82% | 6.71% | $2.50 \times 10^{-19}$ | -0.146 | 0.0179 | $3.39 \times 10^{-16}$ |
| **rs3732218** | Norclozapine | *UGT1A5 UGT1A6 UGT1A7 UGT1A8 UGT1A9 UGT1A10* | A | G | 9.99% | 10.77% | $9.36 \times 10^{-13}$ | -0.078 | 0.0177 | $1.02 \times 10^{-5}$ |
| **rs2472297** | Norclozapine | *CYP1A1 CYP1A2* | T | C | 23.91% | 100% | $2.10 \times 10^{-9}$ | -0.055 | 0.0128 | $1.57 \times 10^{-5}$ |
| **rs1902932** | Ratio | *UGT2B10* | A | G | 9.84% | 40.12% | $1.32 \times 10^{-123}$ | 0.170 | 0.0089 | $4.92 \times 10^{-81}$ |
| **rs115619871*** | Ratio | *UGT2B10* | T | C | 9.82% | 36.88% | $2.30 \times 10^{-123}$ | 0.170 | 0.0089 | $8.25 \times 10^{-81}$ |
| **rs76413136** | Ratio | *CYP2C18* | T | C | 23.25% | 10.60% | $9.18 \times 10^{-22}$ | 0.059 | 0.0065 | $2.29 \times 10^{-19}$ |
| **rs41301394** | Ratio | *POR* | T | C | 26.79% | 43.68% | $4.81 \times 10^{-8}$ | -0.034 | 0.0061 | $2.24 \times 10^{-8}$ |

* Putatively shared causal variant for norclozapine and the metabolic ratio reported by flashfm.

#### Supplementary Figure 1

Manhattan plots of the GWAS analyses of clozapine metabolism carried out in the European subset of CLOZUK. Black horizontal line indicates the genome-wide significant p-value cutoff of $5x10^{-8}$. A: clozapine plasma concentrations ($\lambda_{GC}$= 1.015). B: norclozapine plasma concentrations ($\lambda_{GC}$= 1.017). C: clozapine:norclozapine metabolic ratio ($\lambda_{GC}$= 0.995).

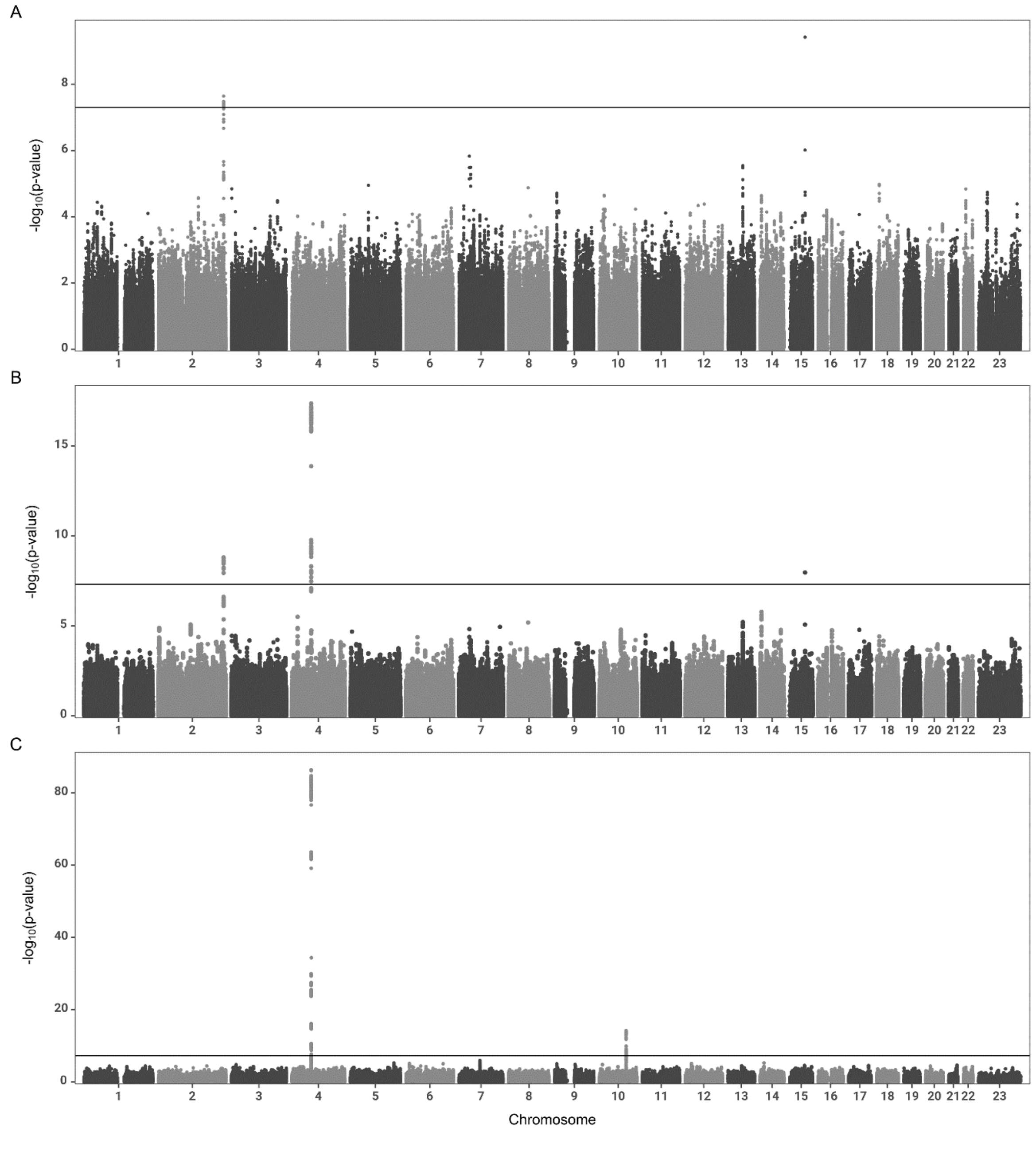

#### Supplementary Figure 2

Manhattan plots of the GWAS analyses of clozapine metabolism carried out in the Sub-Saharan African subset of CLOZUK. Black horizontal line indicates the genome-wide significant p-value cutoff of $5x10^{-8}$. A: clozapine plasma concentrations ($\lambda_{GC}$= 1.066). B: norclozapine plasma concentrations ($\lambda_{GC}$= 1.066). C: clozapine:norclozapine metabolic ratio ($\lambda_{GC}$= 1.082).

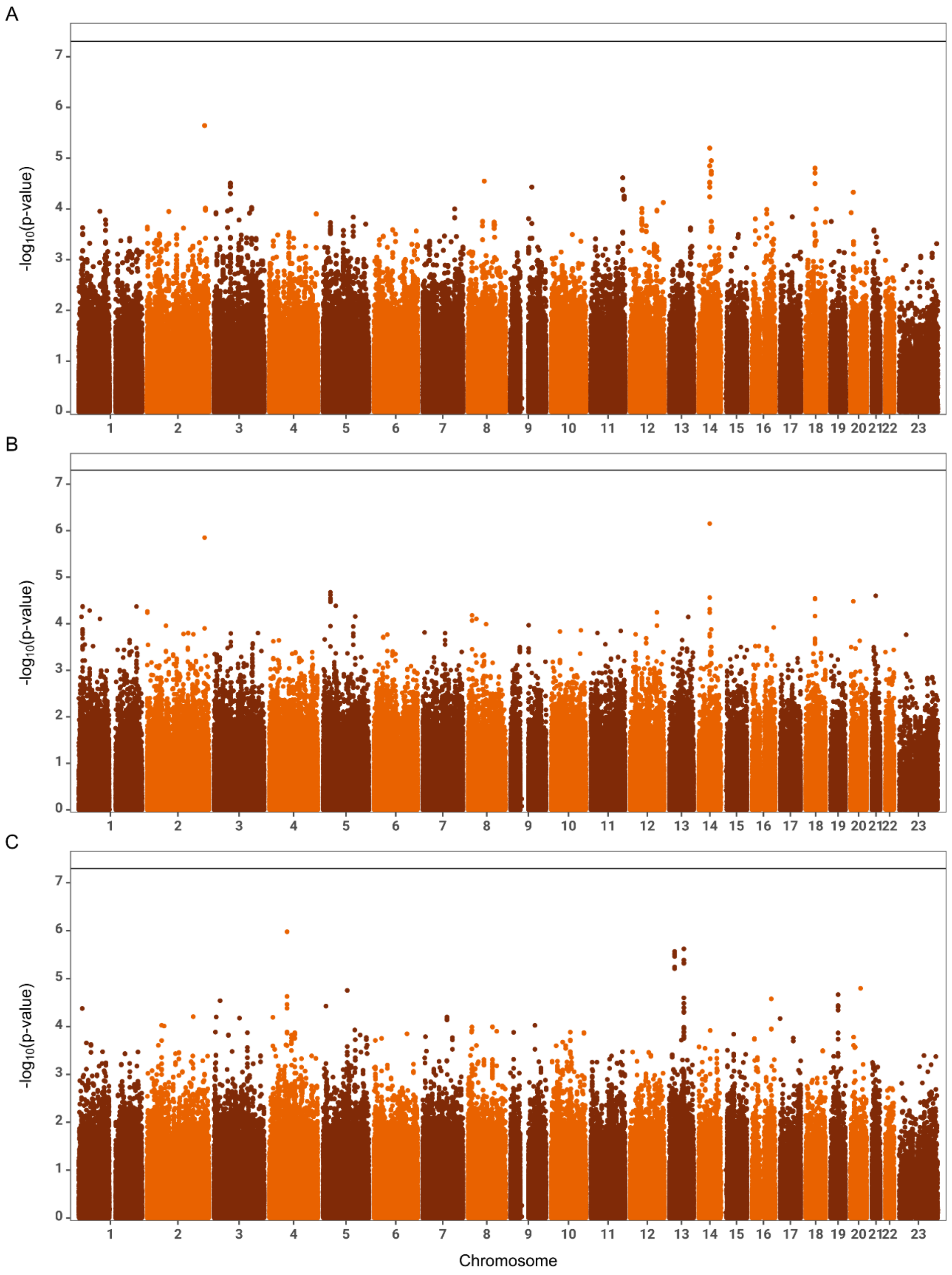

#### Supplementary Figure 3

Manhattan plots of the GWAS analyses of clozapine metabolism carried out in the North African subset of CLOZUK. Black horizontal line indicates the genome-wide significant p-value cutoff of $5x10^{-8}$. A: clozapine plasma concentrations ($\lambda_{GC}$= 1.171). B: norclozapine plasma concentrations ($\lambda_{GC}$= 1.072). C: clozapine:norclozapine metabolic ratio ($\lambda_{GC}$= 1.031).

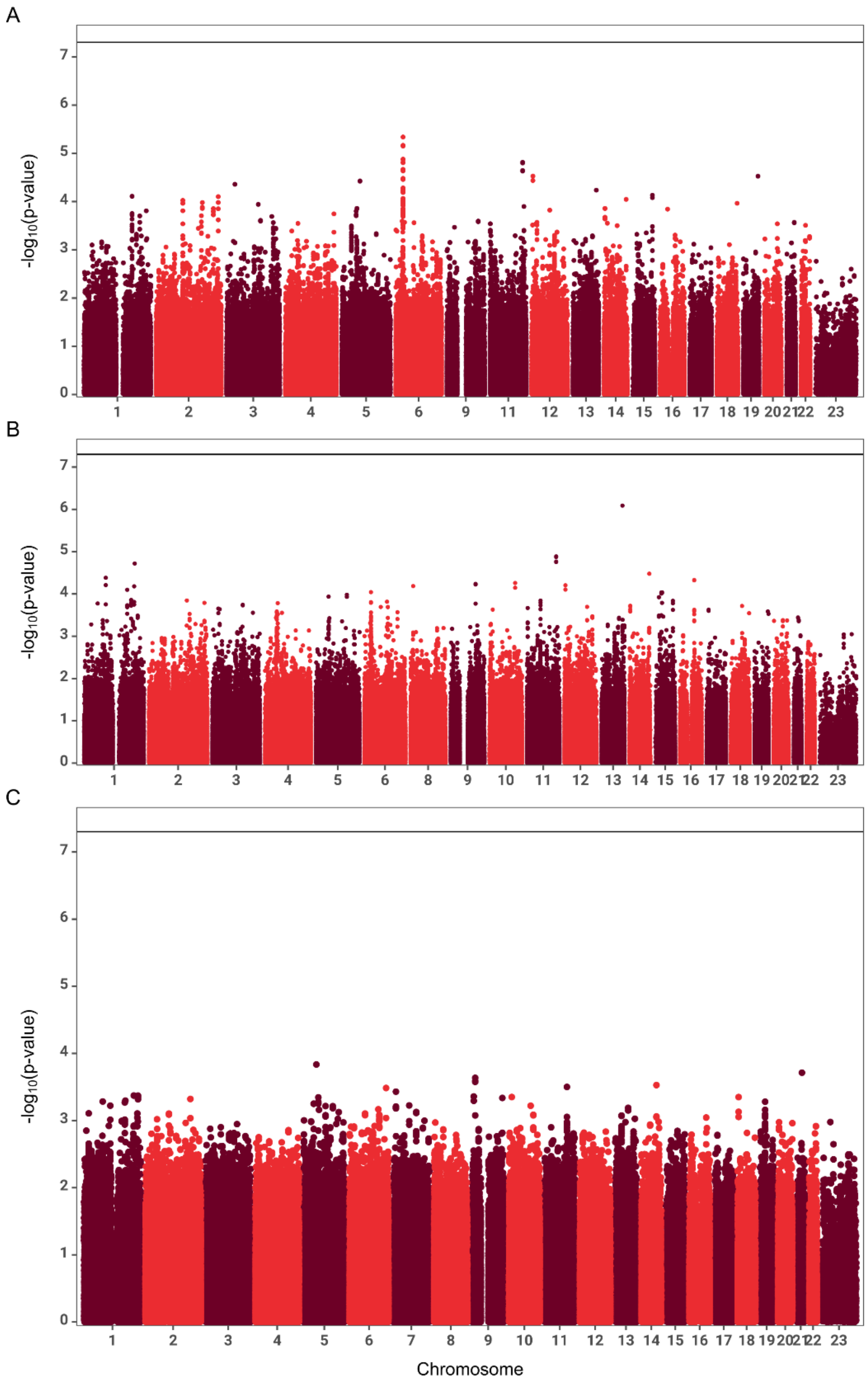

#### Supplementary Figure 4

Manhattan plots of the GWAS analyses of clozapine metabolism carried out in the European subset of CLOZUK. Black horizontal line indicates the genome-wide significant p-value cutoff of $5x10^{-8}$. A: clozapine plasma concentrations ($\lambda_{GC}$= 1.073). B: norclozapine plasma concentrations ($\lambda_{GC}$= 1.076). C: clozapine:norclozapine metabolic ratio ($\lambda_{GC}$= 1.084).

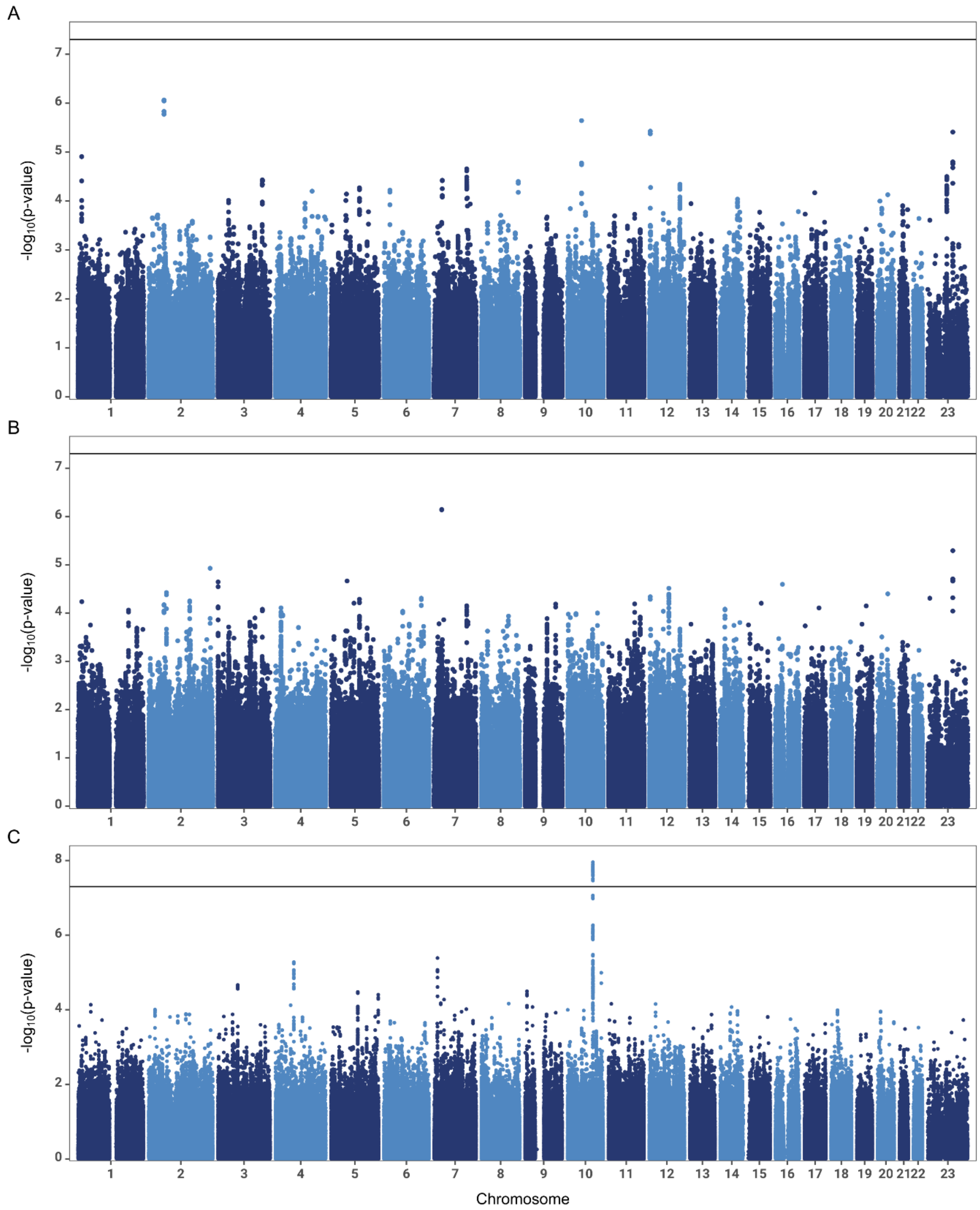

#### Supplementary Figure 5

Association between polygenic scores for clozapine metabolism generated in CLOZUK2 and their corresponding phenotypes in the Sub-Saharan African and Southwest Asian subsets of CLOZUK3. Dots and whiskers indicate the estimated value of the regression effect size and its corresponding 95% confidence interval. Bold colours indicate statistically significant associations; semi-transparent colours indicate non-significant effect sizes.

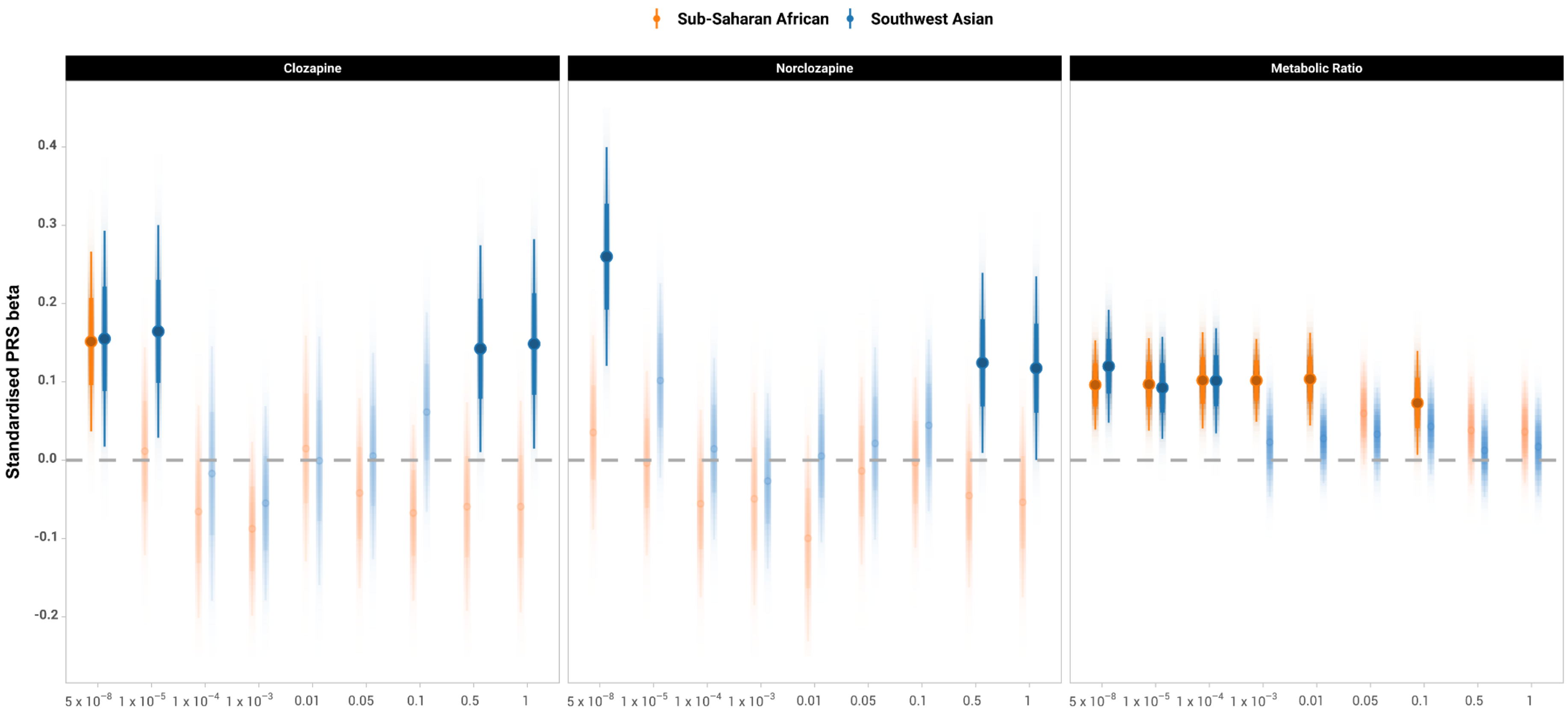

#### Supplementary Figure 6

Full version of **Figure 1C**. Marginal effects of the ancestry groups in the relationship between clozapine doses and the probability of reaching clozapine levels inside or outside the therapeutic range (350-600 ng/mL). Shaded areas in the probability lines highlight a 95% confidence interval. Vertical dashed bars highlight the doses required by individuals in each ancestry group to reach the therapeutic range with 50% probability.

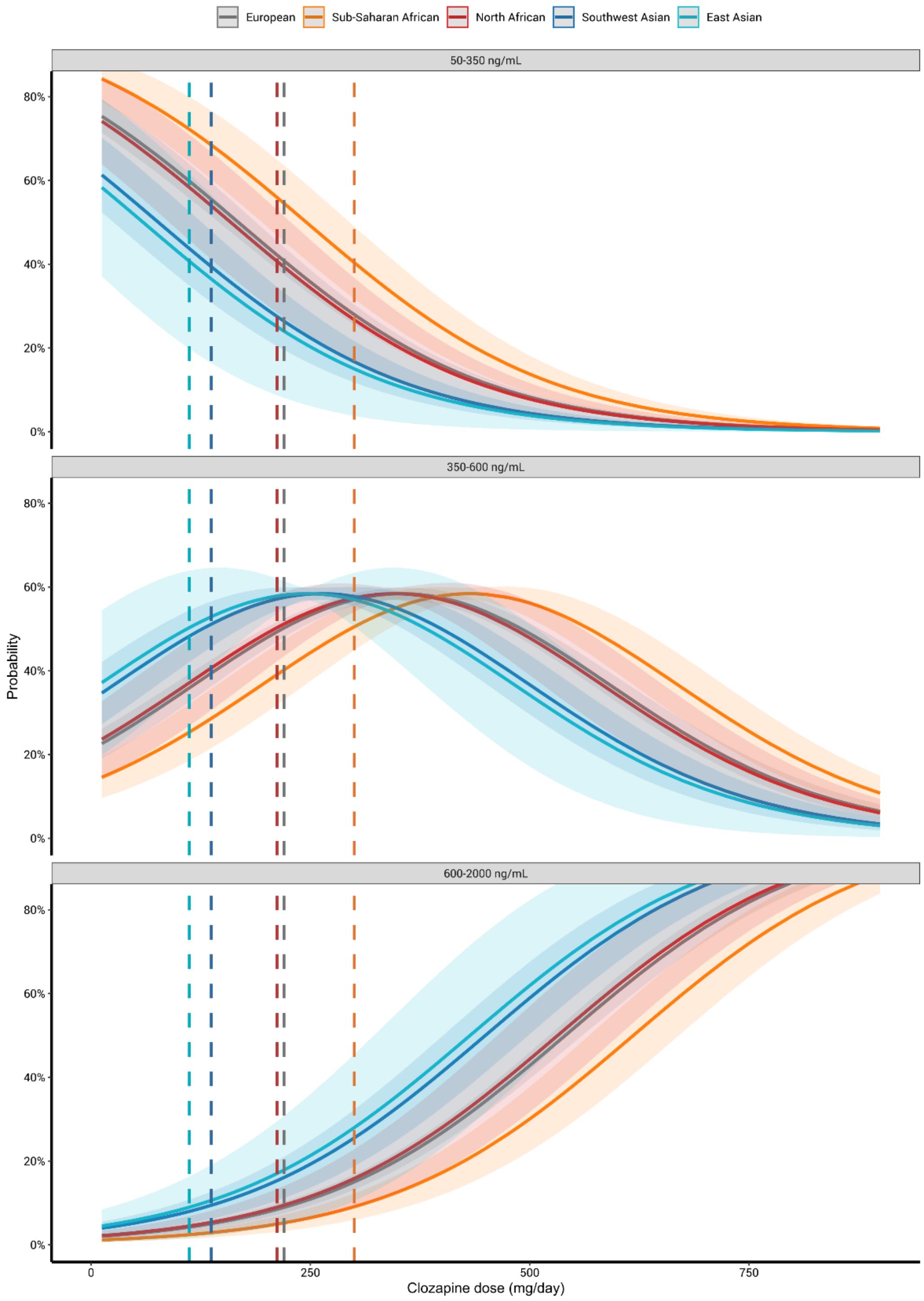

Chang, Christopher C, Carson C Chow, Laurent CAM Tellier, Shashaank Vattikuti, Shaun M Purcell, and James J Lee. 2015. 'Second-generation PLINK: rising to the challenge of larger and richer datasets', *GigaScience*, 4.

Diaz, Francisco J., Jose de Leon, Richard C. Josiassen, Thomas B. Cooper, and George M. Simpson. 2005. 'Plasma clozapine concentration coefficients of variation in a long-term study', *Schizophrenia Research*, 72: 131-35.

Ellison, Justin C., and Robert L. Dufresne. 2015. 'A review of the clinical utility of serum clozapine and norclozapine levels', *Mental Health Clinician*, 5: 68-73.

Flanagan, R J. 2010. 'A practical approach to clozapine therapeutic drug monitoring', *CHMP Bulletin*, 2: 4-5.

Flanagan, R J, J Lally, S Gee, R Lyon, and S Every-Palmer. 2020. 'Clozapine in the treatment of refractory schizophrenia: a practical guide for healthcare professionals', *British Medical Bulletin*, 135: 73-89.

Frei, Oleksandr, Dominic Holland, Olav B. Smeland, Alexey A. Shadrin, Chun Chieh Fan, Steffen Maeland, Kevin S. O'Connell, Yunpeng Wang, Srdjan Djurovic, Wesley K. Thompson, Ole A. Andreassen, and Anders M. Dale. 2019. 'Bivariate causal mixture model quantifies polygenic overlap between complex traits beyond genetic correlation', *Nature communications*, 10: 2417.

Gelman, Andrew, Jennifer Hill, and Aki Vehtari. 2020. *Regression and Other Stories* (Cambridge University Press: Cambridge).

German, Christopher A., Janet S. Sinsheimer, Jin Zhou, and Hua Zhou. 2021. 'WiSER: Robust and scalable estimation and inference of within-subject variances from intensive longitudinal data', *Biometrics*, [in press].

Hernández, N., J. Soenksen, P. Newcombe, M. Sandhu, I. Barroso, C. Wallace, and J. L. Asimit. 2021. 'The flashfm approach for fine-mapping multiple quantitative traits', *Nature communications*, 12: 6147.

Hubbard, Leon, Amy J. Lynham, Sarah Knott, Jack F. G. Underwood, Richard Anney, Jonathan I. Bisson, Marianne.B.M van den Bree, Nick Craddock, Michael O'Donovan, Ian Jones, George Kirov, Kate Langley, Joanna Martin, Frances Rice, Neil Roberts, Anita Thapar, Michael J. Owen, Jeremy Hall, Antonio F. Pardiñas, and James T.R. Walters. 2022. 'DRAGON-Data: A platform and protocol for integrating genomic and phenotypic data across large psychiatric cohorts', *medRxiv*: 2022.01.18.22269463.

Huddart, Rachel, Alison E. Fohner, Michelle Whirl-Carrillo, Genevieve L. Wojcik, Christopher R. Gignoux, Alice B. Popejoy, Carlos D. Bustamante, Russ B. Altman, and Teri E. Klein. 2019. 'Standardized Biogeographic Grouping System for Annotating Populations in Pharmacogenetic Research', *Clinical Pharmacology and Therapeutics*, 105: 1256-62.

Ko, Seyoon, Christopher A. German, Aubrey Jensen, Judong Shen, Anran Wang, Devan V. Mehrotra, Yan V. Sun, Janet S. Sinsheimer, Hua Zhou, and Jin J. Zhou. 2022. 'GWAS of longitudinal trajectories at biobank scale', *The American Journal of Human Genetics*, 109: 433-45.

Legge, Sophie E., Antonio F. Pardiñas, Marinka Helthuis, John A. Jansen, Karel Jollie, Steven Knapper, James H. MacCabe, Dan Rujescu, David A. Collier, Michael C. O'Donovan, Michael J. Owen, and James T. R. Walters. 2019. 'A genome-wide association study in individuals of African ancestry reveals the importance of the Duffy-null genotype in the assessment of clozapine-related neutropenia', *Molecular Psychiatry*, 24: 328-37.

Li, Jun Z., Devin M. Absher, Hua Tang, Audrey M. Southwick, Amanda M. Casto, Sohini Ramachandran, Howard M. Cann, Gregory S. Barsh, Marcus Feldman, Luigi L. Cavalli-Sforza, and Richard M. Myers. 2008. 'Worldwide Human Relationships Inferred from Genome-Wide Patterns of Variation', *Science*, 319: 1100-04.

Lindsey, J. K., B. Jones, and P. Jarvis. 2001. 'Some statistical issues in modelling pharmacokinetic data', *Statistics in Medicine*, 20: 2775-83.

Ma, Clement, Tom Blackwell, Michael Boehnke, and Laura J. Scott. 2013. 'Recommended Joint and Meta-Analysis Strategies for Case-Control Association Testing of Single Low-Count Variants', *Genetic Epidemiology*, 37: 539-50.

McCarthy, Shane, Sayantan Das, Warren Kretzschmar, Olivier Delaneau, Andrew R. Wood, Alexander Teumer, Hyun Min Kang, Christian Fuchsberger, Petr Danecek, Kevin Sharp, Yang Luo, Carlo Sidore, Alan Kwong, Nicholas Timpson, Seppo Koskinen, Scott Vrieze, Laura J. Scott, He Zhang, Anubha Mahajan, Jan Veldink, Ulrike Peters, Carlos Pato, Cornelia M. van Duijn, Christopher E. Gillies, Ilaria Gandin, Massimo Mezzavilla, Arthur Gilly, Massimiliano Cocca, Michela Traglia, Andrea Angius, Jeffrey C. Barrett, Dorrett Boomsma, Kari Branham, Gerome Breen, Chad M. Brummett, Fabio Busonero, Harry Campbell, Andrew Chan, Sai Chen, Emily Chew, Francis S. Collins, Laura J. Corbin, George Davey Smith, George Dedoussis, Marcus Dorr, Aliki-Eleni Farmaki, Luigi Ferrucci, Lukas Forer, Ross M. Fraser, Stacey Gabriel, Shawn Levy, Leif Groop, Tabitha Harrison, Andrew Hattersley, Oddgeir L. Holmen, Kristian Hveem, Matthias Kretzler, James C. Lee, Matt McGue, Thomas Meitinger, David Melzer, Josine L. Min, Karen L. Mohlke, John B. Vincent, Matthias Nauck, Deborah Nickerson, Aarno Palotie, Michele Pato, Nicola Pirastu, Melvin McInnis, J. Brent Richards, Cinzia Sala, Veikko Salomaa, David Schlessinger, Sebastian Schoenherr, P. Eline Slagboom, Kerrin Small, Timothy Spector, Dwight Stambolian, Marcus Tuke, Jaakko Tuomilehto, Leonard H. Van den Berg, Wouter Van Rheenen, Uwe Volker, Cisca Wijmenga, Daniela Toniolo, Eleftheria Zeggini, Paolo Gasparini, Matthew G. Sampson, James F. Wilson, Timothy Frayling, Paul I. W. de Bakker, Morris A. Swertz, Steven McCarroll, Charles Kooperberg, Annelot Dekker, David Altshuler, Cristen Willer, William Iacono, Samuli Ripatti, Nicole Soranzo, Klaudia Walter, Anand Swaroop, Francesco Cucca, Carl A. Anderson, Richard M. Myers, Michael Boehnke, Mark I. McCarthy, Richard Durbin, and Consortium the Haplotype Reference. 2016. 'A reference panel of 64,976 haplotypes for genotype imputation', *Nature Genetics*, 48: 1279-83.

Pardiñas, Antonio F., Mariana Nalmpanti, Andrew J. Pocklington, Sophie E. Legge, Christopher Medway, Adrian King, John Jansen, Marinka Helthuis, Stanley Zammit, James MacCabe, Michael J. Owen, Michael C. O'Donovan, and James T. R. Walters. 2019. 'Pharmacogenomic Variants and Drug Interactions Identified Through the Genetic Analysis of Clozapine Metabolism', *American Journal of Psychiatry*, 176: 477-86.

Paria, Soumya Subhra, Sarthok Rasique Rahman, and Kaustubh Adhikari. 2022. 'fastman: A fast algorithm for visualizing GWAS results using Manhattan and Q-Q plots', *bioRxiv*: 2022.04.19.488738.

Ray, Debashree, and Nilanjan Chatterjee. 2020. 'Effect of non-normality and low count variants on cross-phenotype association tests in GWAS', *European Journal of Human Genetics*, 28: 300-12.

Rights, Jason D., and Sonya K. Sterba. 2020. 'New Recommendations on the Use of R-Squared Differences in Multilevel Model Comparisons', *Multivariate Behavioral Research*, 55: 568-99.

Smith, Robert Løvsletten, Kevin O'Connell, Lavinia Athanasiu, Srdjan Djurovic, Marianne Kristiansen Kringen, Ole A. Andreassen, and Espen Molden. 2020. 'Identification of a novel polymorphism associated with reduced clozapine concentration in schizophrenia patients—a genome-wide association study adjusting for smoking habits', *Translational Psychiatry*, 10: 198.

Stoffel, Martin A., Shinichi Nakagawa, and Holger Schielzeth. 2021. 'partR2: partitioning R2 in generalized linear mixed models', *PeerJ*, 9:e11414.

Terrell, George R. 2003. 'The Wilson-Hilferty Transformation Is Locally Saddlepoint', *Biometrika*, 90: 445-53.

Wilson, Edwin B., and Margaret M. Hilferty. 1931. 'The Distribution of Chi-Square', *Proceedings of the National Academy of Sciences*, 17: 684-88.
